# Investigating the changing taxonomy and antimicrobial resistance of bacteria isolated from door handles in a new infectious disease ward pre- and post-patient admittance

**DOI:** 10.1101/2024.02.02.24302185

**Authors:** Gavin Ackers-Johnson, Ralfh Pulmones, Danielle McLaughlan, Amy Doyle, Joseph M Lewis, Tim Neal, Stacy Todd, Adam P. Roberts

## Abstract

**Background:** Healthcare associated infections (HAIs) are a significant burden to health systems, conferring increased morbidity, mortality and financial costs to hospital admission. Antimicrobial resistance (AMR) further compounds the issue as viable treatment options are constrained. The hospital environment plays a significant role in the development of HAIs, with effective microbial monitoring providing the foundation for targeted interventions and improved infection prevention and control strategies.

**Methods:** This project sampled door handles in an infectious disease ward at the newly built Royal Liverpool University Hospital, Liverpool, UK. Sampling was performed prior to the first patients being admitted to the ward and then six and twelve months after this date. In addition to identifying all isolates, we also investigated the phenotypic antibiotic resistance of all *Staphylococcus* spp. identified, with further whole genome sequencing analysis of multidrug resistant isolates.

**Results:** Prior to patient admission, the majority of isolates identified (57%) were *Staphylococcus* spp., reducing to 32% and 29% at six and twelve months respectively where *Bacillus* spp. accounted for 51% of isolates. No ESKAPE pathogens were identified. Antibiotic susceptibility testing of the *Staphylococcus* spp. showed the rates of resistance were relatively low for all isolates prior to patient admittance, with the exception of cefoxitin (56%). Overall, resistance was highest after six months of ward use, with only tetracycline showing a consistent increase in resistance at each consecutive time point. Despite an increase in isolates susceptible to all antibiotics after 12 months, the rate of multi-drug resistance remained high. Whole genome sequencing revealed the most abundant resistance genes present amongst multidrug resistant staphylococci were *blaZ* (25/26), *mecA* (22/26) and *aac6-aph2* (20/26) respectively, followed by *ermC* (15/26) which was identified in all *Staphylococcus hominis* isolates and *dfrC* (11/26) which was identified in all *Staphylococcus epidermidis isolates*. No isolates believed to be clonal were observed across the three time points assessed.

**Summary:** This study highlighted the prevalence of a resistant reservoir of bacteria recoverable on high touch surfaces. However, given all the isolates identified were unique, it would suggest that the cleaning protocols in place are sufficient, and that the observed bacteria are a result of subsequent recolonisation events. This study emphasises the importance of frequent cleaning and efficient, ongoing, environmental surveillance.

## Introduction

Healthcare associated infections (HAIs) are a significant burden to health systems, and can affect patients, visitors and healthcare workers. The World Health Organisation estimates out of every 100 patients in acute-care hospitals, seven patients in high-income countries and 15 patients in low- and middle-income countries will acquire at least one HAI during their hospital stay (World Health Organization, 2022). Not only are patients faced with poor outcomes in terms of morbidity and mortality, but healthcare providers are faced with increased costs as a result of ongoing treatment and increased patient length of stay (Stewart et al., 2021). The hospital environment plays a significant role in HAIs, where inanimate surfaces may act as a reservoir for pathogens. Admitting a new patient to a room where the previous occupant was infected and/or colonised with a specific pathogen is a risk factor for further transmission (Mitchell et al., 2015). Likewise, cleaning interventions (including chemical, mechanical and human factors) targeted at reducing HAIs, patient colonisation and environmental bioburden often lead to positive outcomes (Peters et al., 2022).

Microbial monitoring of the hospital environment can be a valuable practice, providing the basis for targeted interventions and improved infection prevention and control (IPC) strategies (Sehulster et al., 2004). Furthermore, in hospital settings, where continuous and increased use of disinfectants and antimicrobial drugs create a selective landscape for resistance, it can provide a useful means to screen the local microbiome for clinically relevant antimicrobial resistance (AMR) (Cason et al., 2022).

AMR is one of the top threats to global public health, with bacterial AMR estimated to be directly responsible for 1.27 million and a contributing factor towards 4.95 million global deaths in 2019 (Murray et al., 2022). This issue extends to healthcare settings where the ESKAPE pathogens (*Enterococcus faecium, Staphylococcus aureus, Klebsiella pneumoniae, Acinetobacter baumannii, Pseudomonas aeruginosa, and Enterobacter spp*.), which pose the highest risk of mortality, are responsible for the majority of HAIs and are frequently associated with multidrug resistance (MDR) (Mulani et al., 2019). In addition to the dangers of the ESKAPE pathogens, less clinically significant bacteria colonising environmental surfaces have the potential to act as AMR reservoirs, with dissemination driven by the transfer of mobile genetic elements between bacteria (Larsson & Flach, 2022). If such elements were to be acquired by a pathogen, the treatment of future infections would become increasingly difficult.

This project investigated the changing taxonomy of bacteria isolated from door handles in a new hospital prior to, and following the admittance of patients. We also investigated the phenotypic and genotypic characteristics of antibiotic resistance of all *Staphylococcus* spp. identified.

## Methods

Sample collection was based at the newly constructed Royal Liverpool University Hospital, United Kingdom on an infectious disease ward. Sampling was facilitated at three time points; one week prior to the ward opening to patients, six months and 12 months after the ward had been opened. The project was conducted in conjunction with LUHFT Infection Prevention and Control team. Only environmental sampling occurred with no patient or staff information recorded. In line with NHS Health Research Authority guidance (Is my study research? (hra-decisiontools.org.uk)) this project was considered to be Health Surveillance rather than Research and hence no ethical approval was needed or sought.

At each time point, 40 sites were sampled consisting of stainless-steel lever door handles and push panels. These were situated on the main corridor and the entrance/exit to single occupancy bedrooms with ensuite bathrooms. Whilst the main corridor sites remained consistent at each time point, variable bedrooms were analysed due to access limitations regarding respectful patient care.

25cm^2^ 3D printed thermoplastic (polylactic acid) templates and cotton swabs pre-moistened with neutralising buffer were used to collect samples, swabbing in four directions across the template (up to down, left to right, top-left to bottom-right, top-right to bottom-left).

Bacteria were recovered in 3ml maximum recovery diluent using a Stomacher 80 Biomaster (Seward, Worthing, United Kingdom) at maximum speed for two minutes, with 500µl of diluent plated onto 5% sheep’s blood agar, followed by a subsequent 48-hour incubation at 37°C.

Morphologically distinct colonies were picked from each plate for 16S rRNA gene sequencing. The primers utilised were:

27F - AGA GTT TGA TCC TGG CTC AG

1492R - GGT TAC CTT GTT ACG ACT T

Species identity was determined utilising the closest sequence match when assessed with BLAST (https://blast.ncbi.nlm.nih.gov/Blast.cgi).

All *Staphylococcus* spp. identified were further assessed utilising disc diffusion susceptibility assays following EUCAST guidelines. The antibiotics tested were cefoxitin (30 µg), ciprofloxacin (5 µg), gentamicin (10 µg), trimethoprim/sulfamethoxazole (1:19, 5 µg), tetracycline (30 µg), erythromycin (15 µg) and clindamycin (2 µg). Isolates resistant to three or more classes of antibiotic were classified as multidrug resistant.

All 26 multidrug resistant *Staphylococcus* spp. were submitted to MicrobesNG (https://microbesng.com/) for paired-end 2 x 250bp NovaSeq 6000 Illumina sequencing with a ≥50x target coverage, followed by adapter trimming using Trimmomatic v0.30 (Bolger et al., 2014) with a sliding window quality score cutoff of Q15. *De novo* assemblies were constructed with SPAdes v3.7 (Bankevich et al., 2012) and contigs < 200bp were removed. Assemblies were also manually assessed using Quast v5.02 (Gurevich et al., 2013), with key quality statistics available in the supplementary material 1. The sequence data for this study has been deposited in NCBI BioProject ID: PRJNA1106471.

Genomes were queried against the SRST2-ARGANNOT database (Gupta et al., 2014; Inouye et al., 2014) using ARIBA v 2.14.6 (Hunt et al., 2017) to identify resistance genes. Plasmid replicons were similarly predicted by querying against the PlasmidFinder database (Carattoli et al., 2014).

Intra-species genome assembly relatedness was estimated by Average Nucleotide Identity using FastANI v1.33 (Jain et al., 2018).

## Results

Prior to the opening of the ward, median (interquartile range) colony forming units per cm^2^ (CFU/cm^2^) across all 40 sites was 0.24 (0 to 2.04), increasing to 3.12 (1.14 to 11.64) six months after opening and 13.8 (2.6 to 34) twelve months after opening. Equally, the total number of morphologically distinct colonies identified increased at each time point, being 47, 87 and 126 respectively (we acknowledge that human error could affect these counts as distinctness is open to individual interpretation); additionally, the number of different species of bacteria identified increased, with a total of 13, 23 and 30 different species identified at each time point respectively.

No ESKAPE pathogens were identified from any of the samples. The most prevalent genus of bacteria identified prior to the arrival of patients was *Staphylococcus*, identified at 17/40 sites (43%, 95% CI 33-52%). At this time, only a single *Bacillus* species was identified (3%, 95% CI 0-6%). However, once the ward was in active use, the number of sites where *Bacillus* was identified sharply increased above that of *Staphylococcus* to 27/40 (68%, 95% CI 58-77%) after 6 months and 34/40 (85%, 95% CI 78-92%) after 12 months. Across the same period, the number of sites where *Staphylococcus* was identified slightly increased to 22/40 (55%, 95% CI 45-65%) and 26/40 (65%, 95% CI 56-74%) respectively. *Staphylococcus* spp. and *Bacillus* spp. were the most prevalent genera of bacteria by a large margin.

The greatest prevalence of antibiotic resistance amongst the *Staphylococcus* spp. identified was six months after the ward had been in use, with the highest prevalence of resistance observed across all antibiotics tested except tetracycline as seen in Figure 2. Prior to the ward opening, there was already varying levels of resistance to all antibiotics tested, with tetracycline being the only one where isolates were 100% (27/27) susceptible. Resistance to cefoxitin was already as high as 56% (15/27) and further increased to 71% (20/28) after 6 months of ward use. However, after 12 months this had reduced to 22% (8/37) of isolates. Whilst other antibiotic resistance rates fell close to those observed at the start of the study after the high peak at 6 months, cefoxitin was the only one that went below the initial rate. Tetracycline was the only antibiotic where resistance increased at each consecutive time point. With the exception of one, all isolates which tested resistant to tetracycline were MDR. Similarly, all isolates displaying resistance to gentamicin or trimethoprim/sulfamethoxazole were MDR.

**Figure 1.**
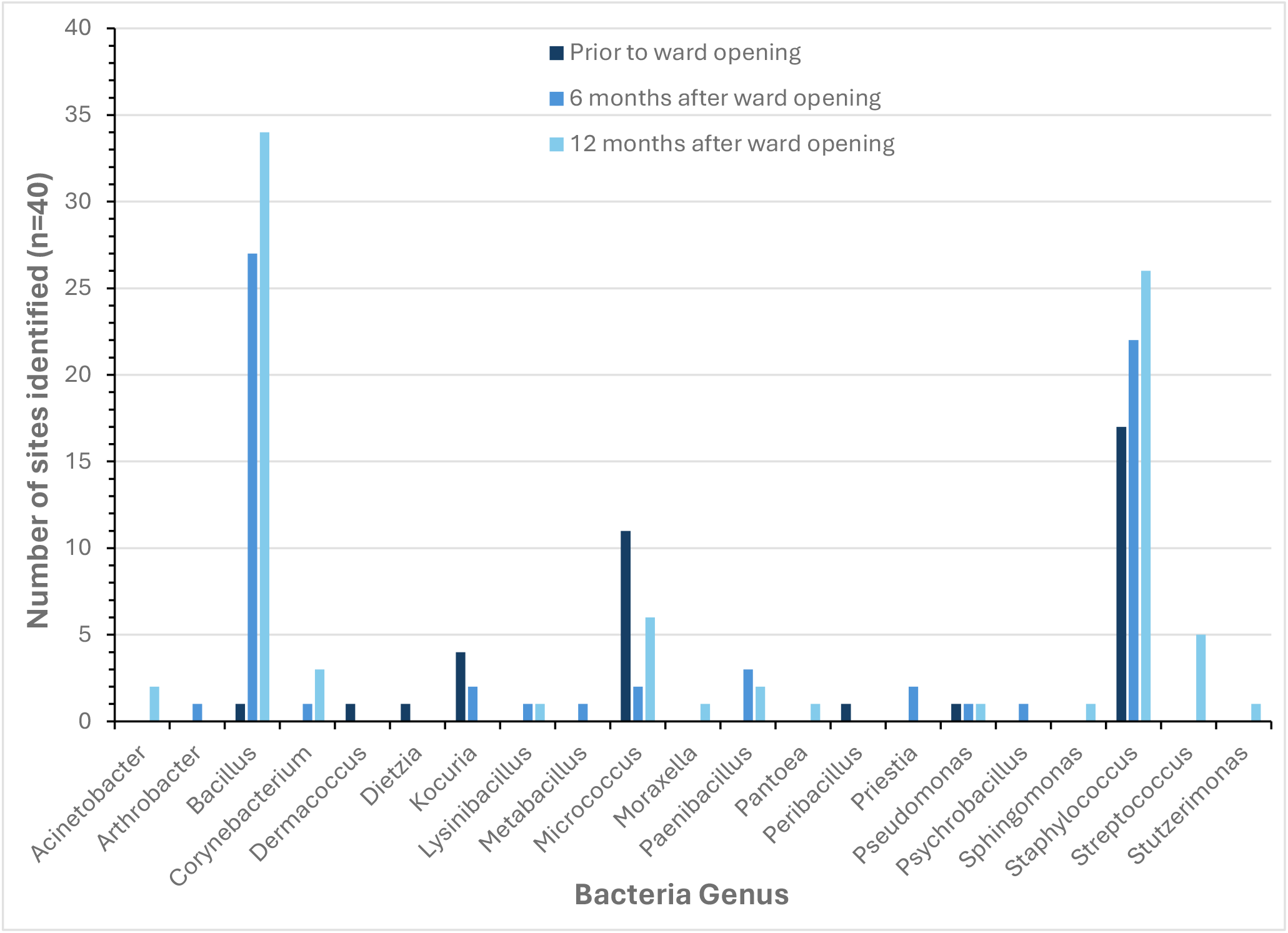
The 16S rRNA gene sequence identity of bacteria isolated from door handles on the infectious disease ward one week prior to, six months after and twelve months after it opened to patients. The data indicates the number of sites the respective genus was identified from a total of 40 sites at each time point.

**Figure 2.**
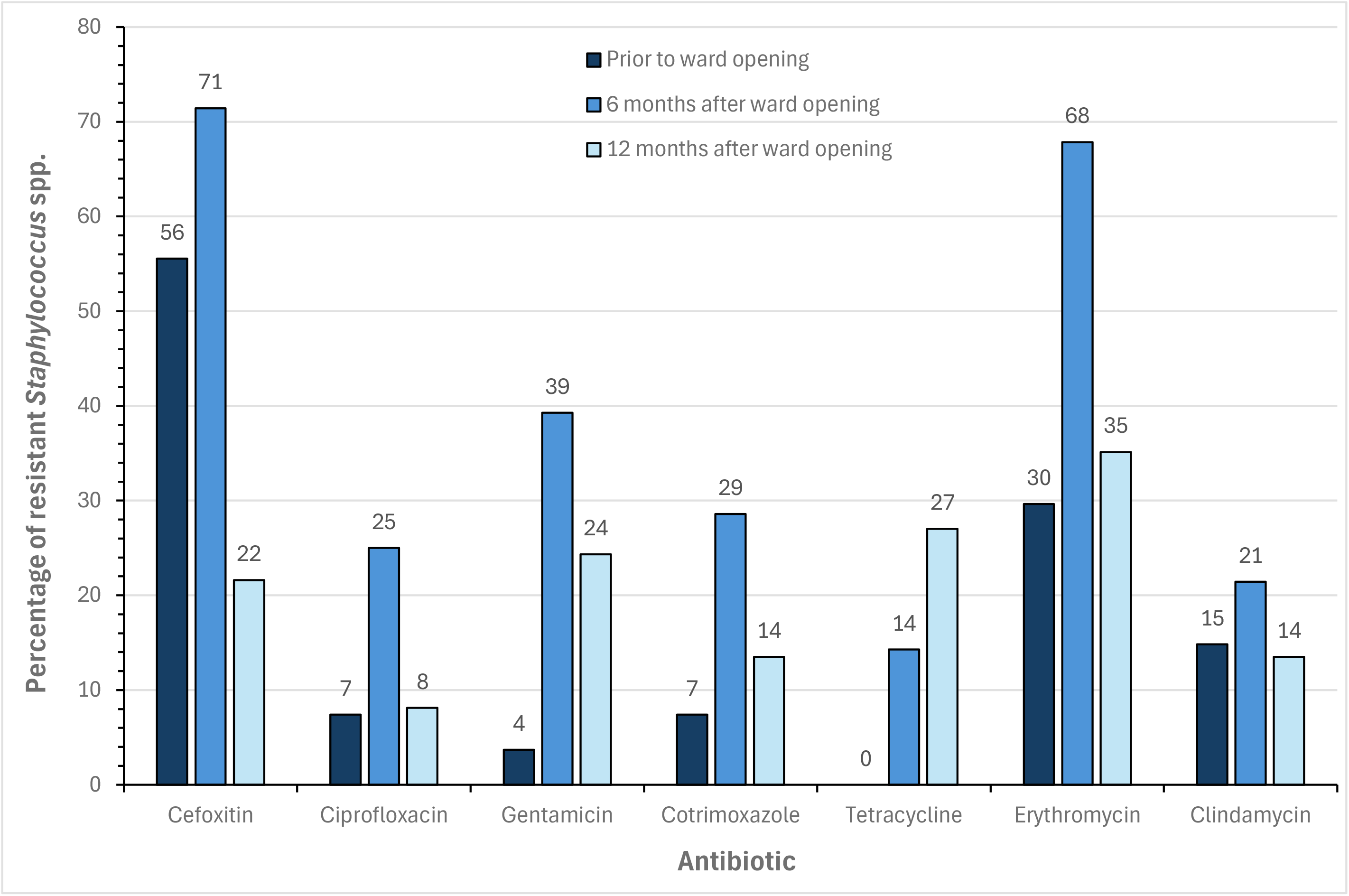
The percentage of *Staphylococcus* spp. resistant to each antibiotic tested at each sample point (prior to ward opening n=27, 6 months after ward opening n=28, 12 months after ward opening n=37).

Whilst overall prevalence of resistance to different agents appear to largely decrease between 6 and 12 months, it is worth noting that the levels of multidrug resistant isolates remain high. Figure 3 shows prior to the opening of the ward, most isolates were either susceptible to all antibiotics tested or resistant to just one. After 12 months of ward use the percentage of isolates susceptible to all antibiotics actually increased relative to the first time point. However, the proportion of multidrug resistant isolates also increased from 7% (2/27) to 27% (10/37) respectively.

**Figure 3.**
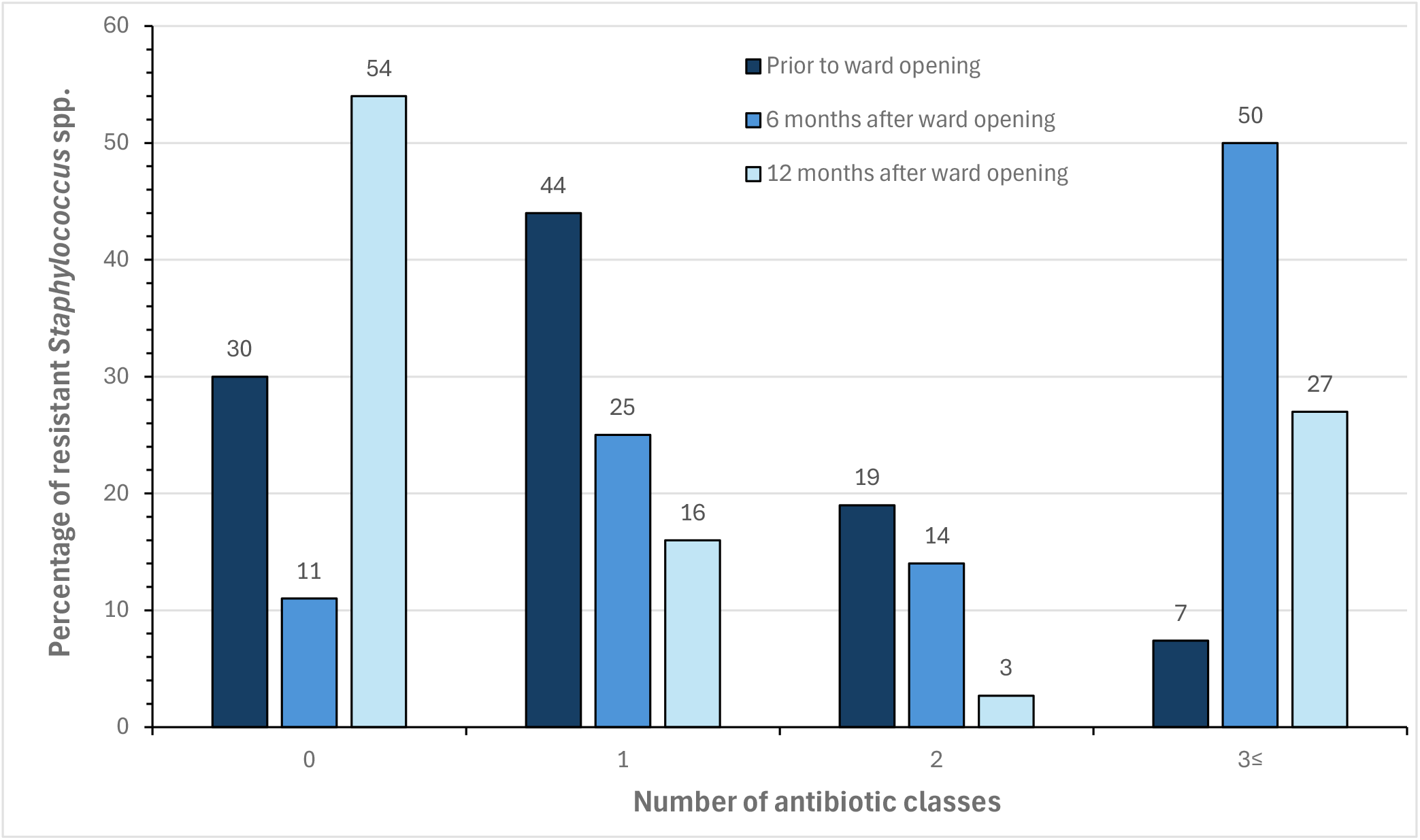
The percentage of *Staphylococcus* spp. identified resistant to 0, 1, 2 or ≥3 different classes of antibiotic (prior to ward opening n=27, 6 months after ward opening n=28, 12 months after ward opening n=37).

Whole genome sequencing analysis of all 26 multidrug resistant *Staphylococcus* spp. (11 *Staphylococcus epidermidis*, 11 *Staphylococcus hominis*, three *Staphylococcus haemolyticus* and one *Staphylococcus capitis*) highlighted the presence of genes and plasmid replicons associated with antimicrobial resistance as seen in Figure 4 below. The genes found at the highest frequency were *blaZ* (25/26), *mecA* (22/26) and *aac6-aph2* (20/26) respectively, followed by *ermC* (15/26) which was identified in all *Staphylococcus hominis* isolates and *dfrC* (11/26) which was identified in all *Staphylococcus epidermidis isolates*. With the exception of trimethoprim/sulfamethoxazole, the associated resistance genes identified were largely in agreement with the observed phenotype. This data is available in supplementary material 2.

**Figure 4.**
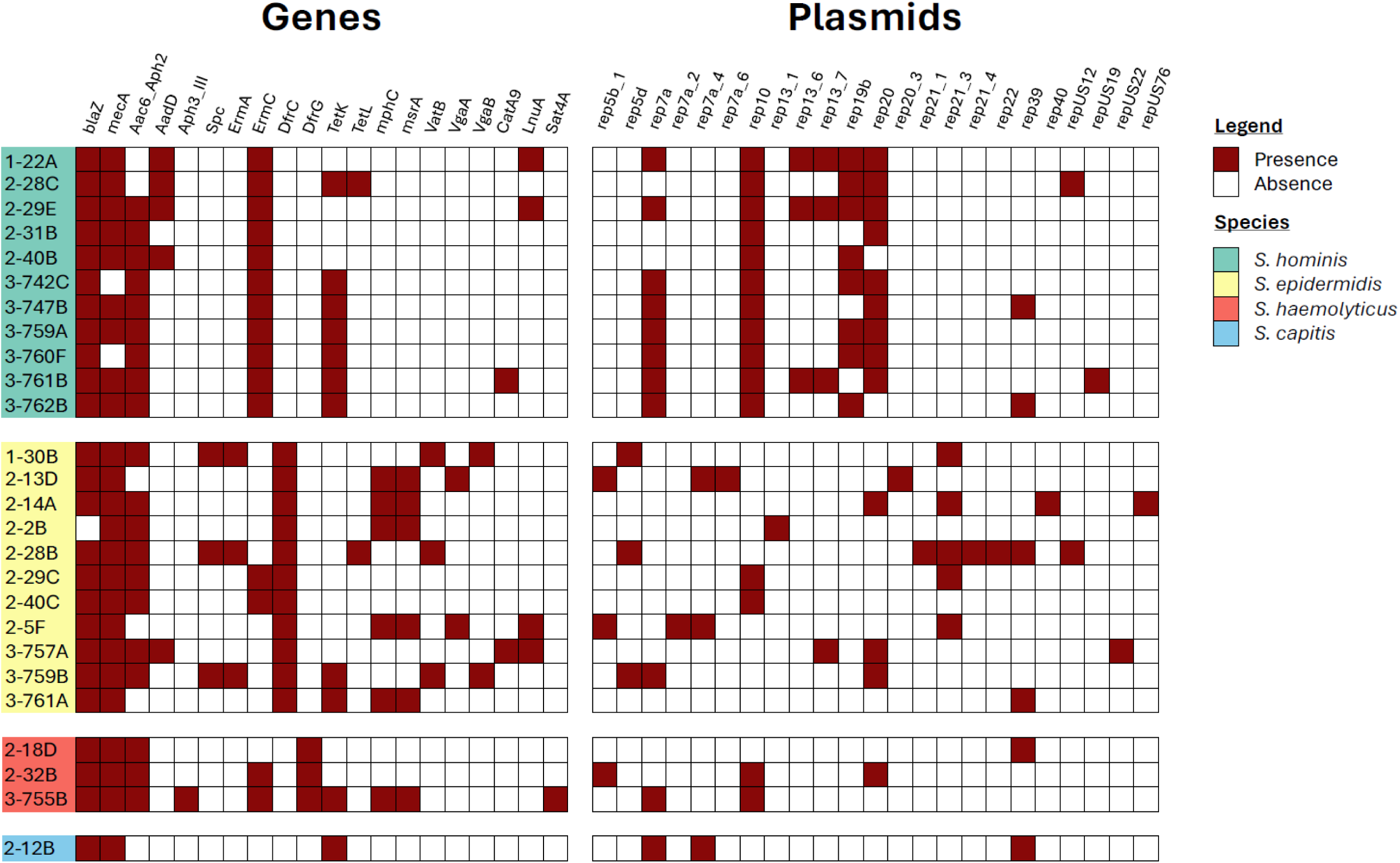
The presence/absence of genes and plasmid replicons associated with antibiotic resistance observed amongst all multidrug resistant *Staphylococcus* spp. collected at three time points; (1-) prior to patient admission, (2-) after six months of ward usage and (3-) after 12 months of ward usage.

There were three *Staphylococcus hominis* isolates (3-747B, 3-759A and 3-762B) and two *Staphylococcus epidermidis* (2-29C and 2-40C) which possessed identical intra-species resistance genes, albeit with varying plasmid replicon profiles, whilst *Staphylococcus hominis* isolates 3-760F and 3-742C possessed both identical resistance genes and plasmid replicons. *Staphylococcus hominis* isolates 1-22A and 2-29E had identical plasmid profiles yet variable resistance gene presence. All remaining isolates had both unique resistance gene and plasmid profiles.

Whilst antimicrobial associated genetic variations were evident across most isolates, high similarities were observed when assessing the intra-species genome assembly relatedness, estimated by Average Nucleotide Identity (ANI) as shown in Figure 5. ANIs ranged from 97.21% - 100%, 99.20% - 99.99% and 99.17% - 99.89% for *Staphylococcus hominis, epidermidis* and *haemolyticus* respectively.

**Figure 5.**
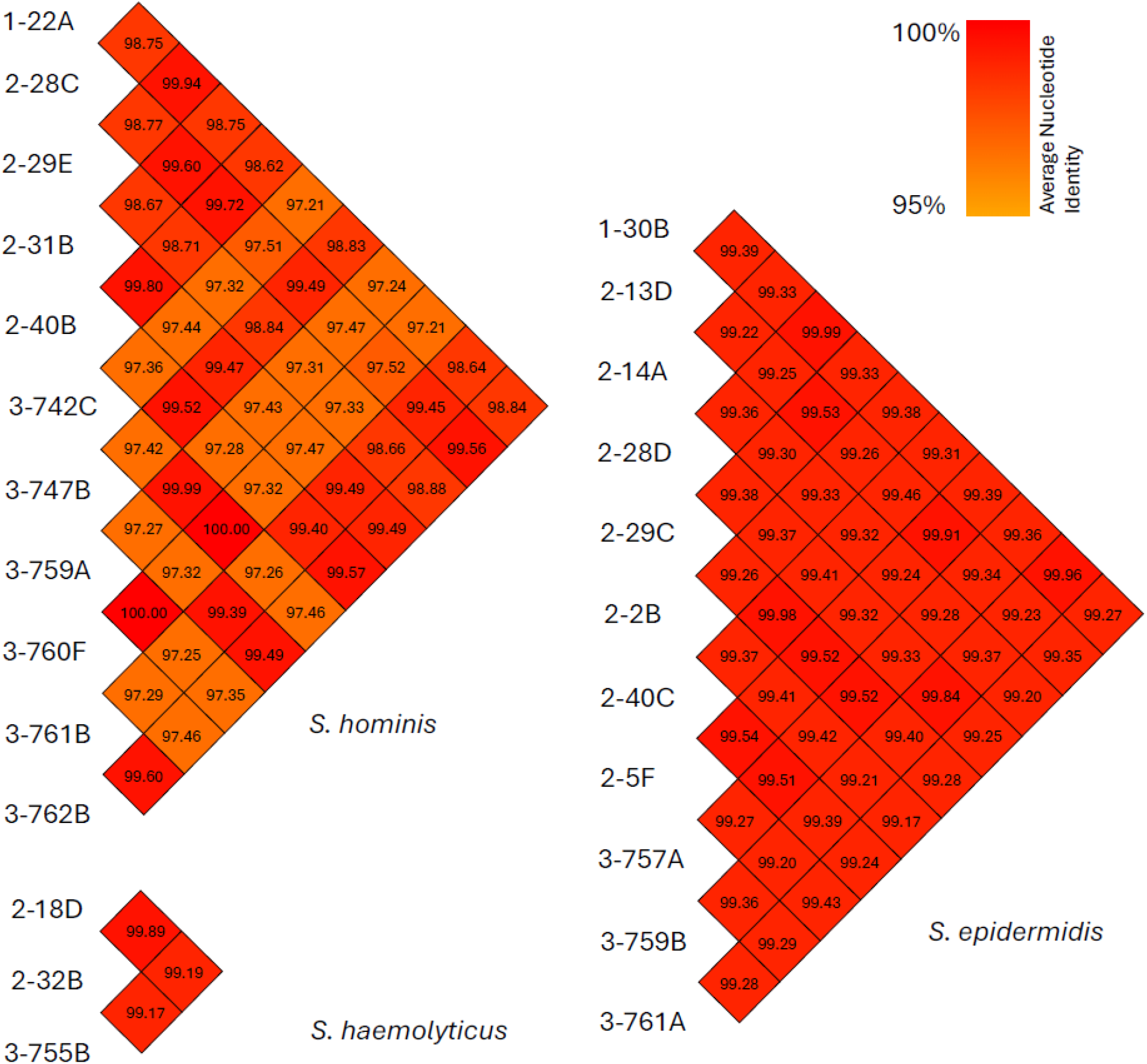
The intra-species genome assembly relatedness of multidrug resistant *Staphylococcus* spp. estimated by Average Nucleotide Identity using FastANI v1.33. *Staphylococcus* spp. were collected at three time points; (1-) prior to patient admission, (2-) after six months of ward usage and (3-) after 12 months of ward usage.

## Discussion

The hospital environment is a known source of bacteria causing nosocomial infection outbreaks (Gastmeier et al., 2006), with healthcare organisations including the UK’s NHS employing a wide array of extensive decontamination protocols in an effort to reduce the environmental bioburden of facilities (Castelli et al., 2022). However, a considerable range of microbial diversity remains (Martineau et al., 2018; Yano et al., 2017). The most clinically significant of these are the ESKAPE pathogens, with third-generation cephalosporin/carbapenem-resistant *Enterobacterales* and carbapenem-resistant *Acinetobacter baumannii* defined by the WHO as “Priority 1: Critical”, and carbapenem-resistant *Pseudomonas aeruginosa*, methicillin-resistant *Staphylococcus aureus* and vancomycin-resistant *Enterococcus faecium* as “Priority 2: High” (Denissen et al., 2022). Within this study, no ESKAPE pathogens (including *Escherichia coli*), MDR or susceptible, were identified. The annual 2022/2023 hospital IPC report does however indicate that at least 51 *E. coli*, 22 *K. pneumoniae*, four *P. aeruginosa*, one methicillin-resistant *Staphylococcus aureus* (MRSA) and 16 methicillin-susceptible *Staphylococcus aureus* (MSSA) hospital onset, hospital associated, infections occurred across LUHFT within a time frame overlapping this study (NHS, 2023). Whilst data collection constrained to three time points could play a role in the lack of ESKAPE pathogens identified, it is likely that other limitations also played a part. Where this project was limited to door handles, previous studies which observed a higher prevalence of priority organisms swabbed a much wider range of environmental surfaces including sinks, tables, bed rails, television remote controls and walls (Anderson et al., 2019; Arduino et al., 2016; Mody et al., 2019; Tanner et al., 2021; van der Schoor et al., 2023). van der Schoor et al. (2023) even noted how nearly all the highly resistant microorganisms they found were present in and around sinks and shower drains as opposed to “dry” surfaces. Furthermore, some of the aforementioned studies utilised broth enrichment, enhancing the detectability of low concentration nosocomial pathogens (French et al., 2011).

*Staphylococcus aureus* is a human commensal organism found on skin and in the nasopharynx, with carriage rates of up to 30% (Wertheim et al., 2005). As such it was anticipated to be found on the door handles sampled within this study. However, this was not the case, as no *Staphylococcus aureus* was identified. This may have been partially influenced by the approach of the hospital to reduce the risk of MRSA infections. As such, the majority of patients are screened for MRSA colonisation either preoperatively or on admission, with positive patients decolonised using standard protocols to reduce the risk of bacteraemia and transmission (NHS, 2023). Whilst this could explain the absence of MRSA, we still would have expected to find MSSA. That being said, multiple *Staphylococcus* species were consistently identified across all time points in relatively high abundance. All of these are known to colonise a specific niche on human skin (Becker et al., 2014), with the exception of *S. pasteuri* which is more closely associated with food specimens (Chesneau et al., 1993). This suggests that microorganisms isolated from door handles are likely derived from human microbiota. Other studies investigating the hospital environment also frequently isolated various *Staphylococcus spp*. (Martineau et al., 2018; Nygren et al., 2023; Wright et al., 2022; Yano et al., 2017), with *S. capitis, S. epidermidis* and *S. hominis* being the most prevalent on frequently touched surfaces (Liu et al., 2022).

Amongst the *Staphylococcus* spp. identified, an initial finding of two MDR isolates, both resistant to cefoxitin, prior to the admittance of patients was noted – without any patients on the ward, these are likely to have originated from healthcare staff or construction workers. This was further compounded by an increase in resistance observed once patients had been admitted. Furthermore, whilst after 12 months the proportion of completely susceptible isolates might have increased (20/37), the isolates resistant to at least one antibiotic were predominantly MDR (10/17), two of which were resistant to all antibiotics tested. Available literature seldom reports on the resistance profiles of coagulase-negative *Staphylococcus* spp. (CoNS) isolated from clinical environments, often focusing on those isolated from clinical cases of infection and/or those colonising healthcare workers. Across these sites there was a consistent observation of high rates of MDR on par with this study (Al-Haqan et al., 2020; Koksal et al., 2009; Ma et al., 2011; Mendes et al., 2010). Similarly, Liu et al. (2022) assessed staphylococci isolated from both hospital personnel and high touch surfaces, observing MDR rates of 61% and 43% respectively. MDR was also prevalent amongst 643 CoNS isolated from a range of non-healthcare associated environmental settings in London, with 6% of isolates fully susceptible, 94% resistant to at least one and 18% resistant to at least 5 antibiotics tested (Xu et al., 2018).

Whilst *S. aureus* is often deemed the most clinically relevant, CoNS are frequently associated with nosocomial infections. In particular, they are known to cause invasive disease in neonates and in the context of immunosuppression or indwelling prosthetic material (Becker et al., 2014). Furthermore, the ability of mobile genetic elements, notably the Staphylococcal cassette chromosome (SCC), to transfer resistance genes amongst *Staphylococcus spp*. provides a pathway for the rapid spread of AMR amongst these opportunistic pathogens, in addition to facilitating the evolution of AMR in *S. aureus* (Cave et al., 2019; Hanssen & Ericson Sollid, 2006; Nasaj et al., 2020). The most prominent resistance gene in this context, the *mecA* gene responsible for methicillin resistance, is a major public health threat (Stefani & Varaldo, 2003). Given its’ significance, resistance to cefoxitin observed within this study of 56% prior to and 71% six months after patient admission appeared high. However, high resistance is frequently seen in clinical isolates, with rates ranging from 57% - 79% (Al-Haqan et al., 2020; Koksal et al., 2009; Ma et al., 2011; Mendes et al., 2010). Furthermore, Liu et al. (2022) found 50% of isolates from healthcare personal and 35% from high touch surfaces were methicillin resistant. These results show that whilst we may have found high levels of cefoxitin resistance, they were in agreement with pre-existing clinical studies, and a figure of 22% after 12 months of ward use was actually much lower than other settings. Given the high prevalence of cefoxitin resistance, including 20/26 MDR isolates, it was to be anticipated that *mecA* would be found in high abundance. Present in 85% (22/26) MDR *Staphylococcus* spp. identified, it correctly predicted phenotypic cefoxitin resistance in 85% (22/26) isolates. Two out of three susceptible isolates with *mecA* present were on the clinical breakpoint susceptibility boundary (22mm), whilst a single isolate was phenotypically resistant despite lacking *mecA*. These observations have been noted before and can be linked to upstream regulatory factors (Marr et al., 2021).

Prior to patient admission, all 27 isolates tested were susceptible to tetracycline; yet six months later, 4/28 (14%) isolates were resistant, all of which were MDR, with one resistant to all antibiotics tested. Again by 12 months, 10/37 (27%) isolates were tetracycline resistant, nine of which were MDR and two of which were resistant to all antibiotics tested. Interestingly, when evaluating the data obtained by Liu et al. (2022), a high proportion of tetracycline resistant isolates were also MDR (8/10 isolates from frequently touched surfaces and 21/23 from healthcare personnel). All phenotypic tetracycline resistance observed amongst the MDR *Staphylococcus* spp. correlated with the presence of *tetK* (11/13) or *tetL* (2/13), both of which encode efflux pumps and are frequently found on small plasmids or, more rarely, integrated into the chromosome or large staphylococci plasmids (Roberts, 1996). These plasmids are mobile and capable of carrying multiple resistance genes, potentially indicating how tetracycline resistance is associated with MDR. As with tetracycline, gentamicin and trimethoprim/sulfamethoxazole resistance was much higher during ward use as opposed to prior to patient admittance, where there was resistance to only a single gentamicin and two trimethoprim/sulfamethoxazole isolates. Equally, all isolates resistant to gentamicin or trimethoprim/sulfamethoxazole were MDR. The presence of *aac6-aph2* correlated closely with gentamicin resistance, with only a single isolate on the breakpoint boundary displaying resistance where the gene was absent. *aac6-aph2* is the only gene currently known to confer gentamicin resistance in *Staphylococcus* and can be located in large plasmids e.g. pSK1 and in chromosomes e.g. SCC*mec* IV (Mlynarczyk-Bonikowska et al., 2022), providing a reasonable basis for the resistance patterns observed.

Conversely, trimethoprim/sulfamethoxazole phenotypic and genotypic resistance correlations had mixed results. *dfrG* was only present in the three *Staphylococcus haemolyticus* isolates, all of which had a matching phenotype. *dfrC* on the other hand, present in all *Staphylococcus epidermidis* isolates and no others, poorly correlated with phenotypic resistance across any of the species. This may be due to dihydrofolate reductase, the enzyme targeted by trimethoprim, having multiple variations spanning across different bacterial species beyond the scope of those analysed (Charpentier & Courvalin, 1997). Trimethoprim/sulfamethoxazole resistance association with MDR is again likely due to the presence of *dfr* genes on transmissible mobile genetic elements (Nurjadi et al., 2014).

The large fluctuations in resistance observed across the study period imply the bacteria present on the sampled hospital door handles is constantly changing and adapting. As indicated by the antibiotic susceptibility data, it would appear as though despite an increase in highly susceptible bacteria by the 12-month time point, a significant MDR cohort of *Staphylococcus* persisted. The average nucleotide identity data corroborated this to an extent, particularly with *Staphylococcus epidermidis*, where percentage similarities were consistently high. However, discussions are ongoing as to how to appropriately classify relationships with respect to ANI values. Typically, a threshold of >95% signifies the same species, >99.5% for the same sequence type and approaching 100% for clonal relationships (Rodriguez-R et al., 2024; Varghese et al., 2015). With these breakpoints in mind, there appears to be multiple cases of highly related sequence types spanning across all three sample points, with a select few potentially clonal relations. The most prominent of these are *Staphylococcus hominis* isolates 3-760F and 3-742C, sharing 100% similarity in terms of ANI, resistance genes and plasmid replicons. These were isolated at the same time point from a bedroom exit and a dirty utility room exit respectively and are likely to be clonal. Isolate 3-759A was isolated from the entrance to the same bedroom as 3-760F. These two isolates also shared 100% ANI, had identical plasmid replicons and near-identical resistance genes, the exception being 3-759A harboured *mecA* where 3-760F did not. As such, it is unlikely that they are clonal unless the *mecA* gene had only just been acquired very recently. Crucially, these high similarities did not extend across different time points. Even amongst *Staphylococcus epidermidis*, isolates 1-30B and 2-28D, collected in the first and second cohort respectively, shared the highest ANI similarity of 99.99%, yet only one has a *tetL* gene and the other a *vgaB*, with further variations in their plasmid replicons. Again, these are unlikely to be clonal. As such, we identified that a persistent cohort of increasingly MDR staphylococci was not present. Instead, we hypothesise the microorganisms recovered from hospital door handles change over time, with cleaning protocols effectively removing and / or killing those present and subsequent recolonisation events leading to the addition of new bacteria.

Whilst *Staphylococcus spp*. formed the predominant genus isolated pre-patient admission, after six and twelve months of ward usage *Bacillus* spp. accounted for 51% of all isolates. This is likely due to their wide distribution in the environment, particularly in soil, and association with food products (Rana et al., 2020). Similar to this study, Al-Habibi et al. (2022) examined 407 environmental isolates across three hospitals, identifying 43.2% as *Bacillus* spp. and 19.2% as CoNS. The *Bacillus* genus has long been considered too broad, with many members being incrementally reclassified (Gupta et al., 2020). Several of these were identified within this study including *Metabacillus, Paenibacillus, Peribacillus, Psychrobacillus*, and *Priesta* species. All of these are frequently found in soil and rarely cause disease (Biedendieck et al., 2021; de Andrade et al., 2023; Krishnamurthi et al., 2010; Sáez-Nieto et al., 2017). The most clinically significant *Bacillus* species identified was *Bacillus cereus*, frequently associated with food-borne outbreaks and more recently implicated in localised wound and eye as well as systemic infections (Bottone, 2010; Glasset et al., 2018). Given its wide prevalence in the environment, it does not provide cause for immediate concern. However, it is something that should be monitored over time. The large majority of other bacteria isolated as part of this study bear little clinical relevance and were observed in agreement with other previous studies, albeit with *Streptococcus* spp. identified at much lower levels (Martineau et al., 2018; Nygren et al., 2023; Yano et al., 2017).

## Conclusion

The presence of a resistant reservoir of bacteria recoverable on high touch surfaces highlights the importance of extensive and sustained cleaning protocols and efficient environmental surveillance systems, especially considering CoNS are being increasingly viewed as emerging pathogens. That being said, unique antimicrobial profiles and a lack of potentially clonal isolates observed across different time points signifies the cleaning protocols in place are effective. This was further justified by the absence of ESKAPE pathogens including *Staphylococcus aureus*. Instead, the presence of the observed bacteria can likely be attributed to post-cleaning recolonisation events.

In future, it would be beneficial to expand such studies to a greater variety of sites in addition to door handles to ensure an accurate representation of the hospital environment and respective microbiome. Furthermore, regularly assessing the bacteria colonising patients and healthcare staff would shed light on potential routes of transmission and recolonisation of high touch surfaces.

## Supporting information

Supplementary material 1 - WGS Assembly Statistics

Supplementary material 2 - AMR phenotype and genotype of Staphylococcus species

## Data Availability

All data produced in the present work are contained in the manuscript.

https://www.ncbi.nlm.nih.gov/bioproject/PRJNA1106471/

